# Renal and systemic hemodynamic effects of empagliflozin: Three randomized, double blind, placebo controlled cross-over trials

**DOI:** 10.1101/2024.05.28.24308079

**Authors:** Steffen Flindt Nielsen, Camilla Lundgreen Duus, Niels Henrik Buus, Jesper Nørgaard Bech, Frank Holden Mose

**Author notes:** Corresponding author: Steffen Flindt Nielsen, Adress: Gødstrup Hospital, Hospitalsparken 15, 7400 Herning, Denmark.

## Abstract

**Background:** Sodium-glucose cotransporter 2 inhibitors (SGLT2i) improve renal outcomes in type 2 diabetes mellitus (DM2) and chronic kidney disease (CKD). A decrease in renal blood flow (RBF) with attenuation of glomerular hyperfiltration may contribute to this. We examined renal and systemic hemodynamic effects of SGLT2i in relevant patient categories.

**Methods:** Using a double-blind placebo controlled cross-over design we randomized patients with DM2 and estimated glomerular filtration rate (eGFR) >60 ml/min/1.73m^2^ (n=16), patients with DM2 and eGFR 20-60 ml/min/1.73m^2^ (n=17) and patients with non-diabetic CKD and eGFR 20-60 ml/min/1.73m^2^ (n=16) to empagliflozin 10 mg daily or placebo for four weeks and crossed over to the opposite treatment after two-week washout. RBF was measured with ^82^Rubidium positron emission tomography/computed tomography (^82^Rb-PET/CT), GFR as plasma clearance of ^99m^Technetium-diethylene-triamine-pentaacetate, while 24-hour blood pressure (BP) and total peripheral vascular resistance (TVR) were recorded using the commercially available Mobil-O-graph.

**Results:** Compared to placebo empagliflozin reduced RBF by 6% in the DM2-CKD group (p<0.001), while there were non-significant decreases of 4% in the DM2 group and 1% in the CKD group (p=0.29 and 0.72). Empagliflozin reduced GFR, BP and TVR in all groups. Although total renal vascular resistance (RVR) remained unchanged, calculations based on Gomez’ equations revealed a reduction of post-glomerular resistance in the DM2 and CKD groups.

**Conclusion:** Short-term empagliflozin treatment reduced RBF in patients with DM2 and CKD, whereas GFR, BP and TVR were reduced in all groups. The lack of reduction in total RVR together with a decrease in post-glomerular resistance and systemic BP suggest SGLT2i protect the glomerulus due to relative pre-glomerular vasoconstriction and post-glomerular vasodilation.

**Registration:** EU Clinical Trials Register 2019-004303-12, 2019-004447-80 and 2019-004467-50

**Clinical Perspective:** *What is new?:*

- This is the first study of the hemodynamic effects of sodium-glucose cotransporter 2 inhibitors in diabetic and non-diabetic chronic kidney disease.
- We found that the sodium-glucose cotransporter 2 inhibitor empagliflozin reduced renal blood flow in patients with type 2 diabetes and chronic kidney disease.
- Empagliflozin reduced blood pressure and total vascular resistance in patients with type 2 diabetes both with and without chronic kidney disease and in patients with non-diabetic chronic kidney disease.

*What are the clinical implications?:*

- This is the first time sodium-glucose cotransporter 2 inhibitors have been shown to decrease renal blood flow in patients with type 2 diabetes, corroborating the hypothesis that they exert clinical benefits through attenuation of hyperfiltration
- Our findings suggest a combined pre- and post-glomerular hemodynamic response that may underlie the beneficial clinical effects.
- The reduction in blood pressure and total peripheral resistance point to a novel vascular effect of empagliflozin that is present in both patients with and without type 2 diabetes or chronic kidney disease.

## Introduction

Since the emergence of sodium-glucose cotransporter 2 inhibitors (SGLT2i) their precise mechanisms of action have been a topic of debate. According to the tubular hypothesis of hyperfiltration, SGLT2i can confer renal protection via changes in renal hemodynamic function [1]. SGLT2 is thought to play a key role in the pathophysiology of diabetic nephropathy, mediating reuptake of excessively filtered sodium and glucose thereby decreasing sodium delivery to the *macula densa*. This triggers a tubulo-glomerular feedback (TGF) response which dilates the afferent arteriole increasing renal blood flow (RBF) and glomerular pressure, leading to hyperfiltration and elevated glomerular filtration rate (GFR). Long term hyperfiltration can lead to glomerular injury and chronic kidney disease (CKD) [2]. Conversely, blockage of SGLT2 reduce sodium and glucose reuptake which increases distal sodium delivery, causing afferent arteriolar vasoconstriction, decreased RBF and alleviation of hyperfiltration, resulting in kidney protection [3–6]. Cherney et al. showed that treatment with empagliflozin reduced effective renal plasma flow (ERPF) and GFR in younger patients with type 1 diabetes (DM1), indicating ameliorated glomerular hyperfiltration [7] and studies in mice have demonstrated that empagliflozin can induce afferent arteriolar vasoconstriction and reduce hyperfiltration at the single nephron level [8].

In patients with DM2 and/or CKD initiation of SGLT2i is associated with an acute, but reversible, reduction in estimated GFR (eGFR) [9] whereafter it stabilizes, mitigating further decline in renal function [10, 11]. This acute eGFR reduction is not related to increased risk of renal events; on the contrary it predicts better outcomes [12, 13]. This seems to corroborate the above hypothesis, however none of the studies examining renal hemodynamic effects of SGLT2i in patients with DM2 have been able to document a decrease in RBF [14–16]. One explanation could be that the hemodynamic effects of SGLT2i differ between patients with DM1 and DM2 with post-glomerular vasodilation being more prominent in the latter [17].

Despite the extensive use of SGLT2i in patients with diabetic and non-diabetic CKD, their hemodynamic effects have not been studied in these groups. Therefore, we examined the renal and systemic hemodynamic effects of empagliflozin in patients with DM2 with and without concomitant CKD as well as in patients with non-diabetic CKD, reflecting the patient categories who are offered SGLT2i treatment. We employed a novel ^82^Rubidium positron emission tomography/computed tomography (82Rb-PET/CT) method for estimating RBF, our primary endpoint. Secondary endpoints included GFR, 24-hour ambulatory blood pressure (BP), total peripheral vascular resistance (TVR) and renal vascular resistance (RVR). We hypothesized that empagliflozin would reduce RBF and decrease GFR and BP in all groups.

## Methods

### Design

We conducted three randomized, double-blind, placebo controlled, single centre cross-over studies including participants with DM2 and preserved kidney function (DM2 group), DM2 and concomitant CKD (DM2-CKD group) and non-diabetic CKD (CKD group). The design and methods of our project, the so called SiRENA project, have been previously published [18]. Here, we report the primary endpoint and selected secondary outcomes. The experimental setup was identical in the three studies (Figure 1). Briefly, participants were randomized to four weeks of treatment with either empagliflozin 10 mg daily or matching placebo and crossed over to four weeks of the opposite treatment after a wash out period of at least 2 weeks. Study endpoints were measured at an examination day at the end of each treatment period. If participants were on an SGLT2i at inclusion, it was discontinued at least two weeks prior to randomization. If necessary, it could be substituted to a different class of anti-diabetic treatment in accordance with national treatment guidelines. All other medications were kept unaltered. Four days prior to each examination, participants were encouraged to adhere to their usual diet and fluid intake was standardized.

**Figure 1:**
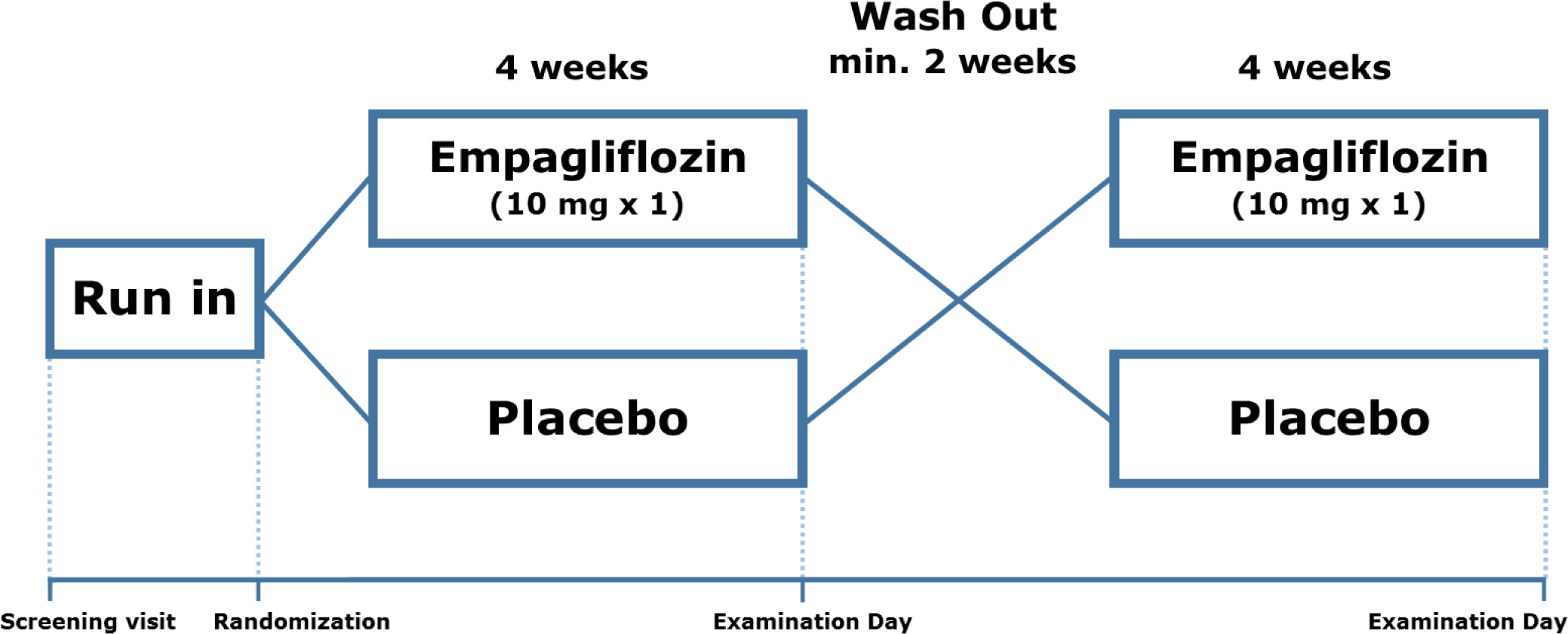
Study design. Schematic view of the study design

### Study participants

All participants were informed of the study purpose and methods and signed consent was obtained before screening. Participants were screened prior to inclusion, ensuring they fulfilled all inclusion and no exclusion criteria. Inclusion criteria were: Age > 18 years, DM2 with a glycated hemoglobin (Hb1Ac) between 48-70 mmol/mol, diagnosed at least one year prior to inclusion and on stable anti-diabetic treatment for at least 3 months (DM2 and DM2-CKD groups) and eGFR > 60 ml/min/1.73m^2^ (DM2-group) or 20-60 ml/min/1.73m^2^ (DM2-CKD and CKD-groups). Fertile women were to use safe contraception. Exclusion criteria were: Anamnestic or clinical signs of heart- or liver failure, active malignancies besides non-melanoma skin-cancers, body mass index (BMI) > 35 kg/m^2^, pregnancy or nursing, alcohol or substance abuse, allergies or unacceptable side effects to the study medication, previous kidney transplantation, autosomal dominant polycystic kidney disease (ADPKD), DM1 (or DM2 for the CKD group) or if participants were deemed unfit to complete the trial. The studies were approved by The Central Denmark Region Committees on Health Research Ethics and the Danish Medicines Agency and were conducted in accordance with the Declaration of Helsinki 2013. The studies were monitored by the Good Clinical Practice (GCP) Unit of Aarhus and Aalborg Universities.

### Statistics and sample size calculation

Normality was evaluated using histograms and QQ-plots. Variance was evaluated using Bland-Altman and scatter plots. If non-normally distributed, normality was tested on the logarithmic scale. Paired t-test was applied to normally distributed data and is presented as mean ± standard deviation (SD) or as geometric mean [inter quartile range (IQR)] if on the log scale. The sign-test was applied to non-parametric data. Potential period and carry-over effects were assessed using a linear mixed effects model with treatment, period and carry-over as fixed effects and participant ID as a random effect. Between-group differences were tested using a linear mixed effects model, allowing for unequal variance. p-values <0.05 were considered statistically significant. A power calculation was performed on the primary endpoint. Fifteen patients were needed in each study to detect a minimal relevant difference in RBF of 0.167 ml/min/ccm, with a SD of 0.180 ml/min/ccm, a two-sided α- level of 5% and a power of 90%. All statistical analyses were done using STATA version 18.0 (StataCorp LLC, College Station, Texas, US).

## Study methods

### RBF

RBF was estimated based on renal uptake of ^82^Rb measured with ^82^Rb-PET/CT scans. Scanning procedure and data analysis was performed as described by Langaa et al. [19, 20]. In short, a low dose CT scan was used for attenuation correction. Following bolus injection of 555 MBq of ^82^Rb, an 8-minute dynamic PET-scan was performed with the abdominal aorta (AA) and both kidneys within the same field of view. Renal ^82^Rb activity was estimated using image based iso-contouring of both kidneys. AA activity was defined by placing a volume of interest (VOI) within the proximal AA. Time activity curves were generated on the basis hereof (see Figure S1). We estimated RBF (ml/min/ccm) for each kidney as the single kidney K_1_-value using a 1-tissue compartment model based on renal ^82^Rb activity uptake with AA activity as input function. Mean RBF was estimated for each participant as the mean K_1_-value. AA measurements were corrected for partial volume and spill-over and all measurements were β-corrected as has been previously described. Analyses were made using PMOD^©^ (PMOD Technologies Ltd., Zürich, Switzerland). Total RBF (ml/min) was assessed by multiplying RBF with kidney volume as estimated by the renal iso-contours using the formula:

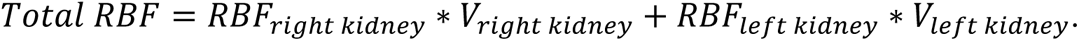

Effective renal plasma flow (ERPF) was calculated as

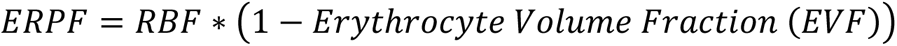

### GFR

GFR was measured by clearance of ^99m^Technetium-diethylene-triamine-pentaacetate (^99m^Tc-DTPA). First, a zero sample was drawn. Hereafter 25 MBq of ^99m^Tc-DTPA was injected intra-venously and blood samples were drawn after three, four and five hours, measuring residual plasma activity. Measurements were standardized for height and weight.

### Systemic hemodynamics

24-hour BP, heart rate and TVR were measured using a Mobil-O-Graph® (I.E.M. GmbH., Aachen, Germany). The Mobil-O-Graph was placed on the upper forearm at the beginning of the examination day and was removed 24 hours later. Measurements were done every 20th minute.

### Renal hemodynamic calculations

Filtration fraction (FF) was calculated as 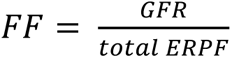. Mean arterial pressure (MAP) was calculated as 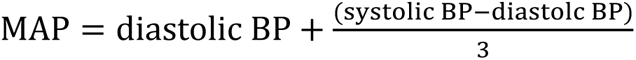, where day-time BP values were used. Renal vascular resistance (RVR) was calculated as 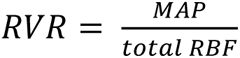. Intra glomerular pressure (P_GLO_), afferent and efferent glomerular arteriolar resistance (R_a_ and R_e_) were estimated using the Gomez equations [21].

### Biochemical analysis

Plasma and urine samples were analysed by the Department of Clinical Biochemistry, Gødstrup Hospital as routine procedures, except for urinary glucose which was measured using an Invitrogen Glucose Colorimetric Detection Kit (ThermoFisher Scientific Inc., Waltham, MA. USA). eGFR was calculated using the CKD-EPI formula. Urinary albumin/creatinine ratio (uACR) was calculated based on measurements of urinary albumin and creatinine from the total voided volume during the examination day (collection time: 5 hours)

## Results

Inclusion began in April 2021 and was completed by September 2022. Examinations were completed by December 2022. 49 participants completed the studies; 16 in the DM2 group, 17 in the DM2-CKD group and 16 in the CKD group. For further elaboration on inclusion and exclusion, see Figure 2. Baseline characteristics are described in Table 1.

**Figure 2:**
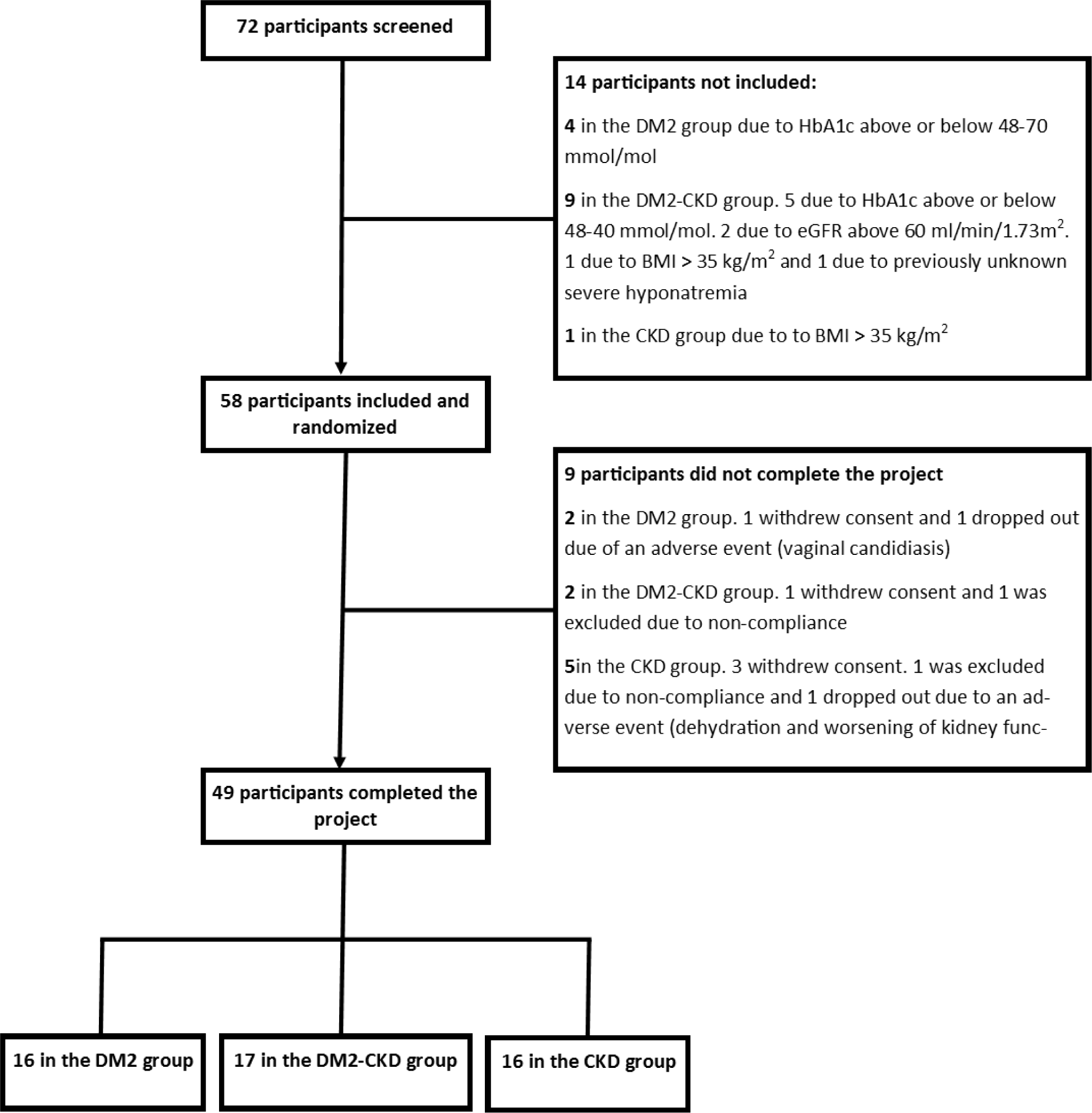
Flow-chart of screening, inclusion, and exclusion for all studies.

**Table 1:**
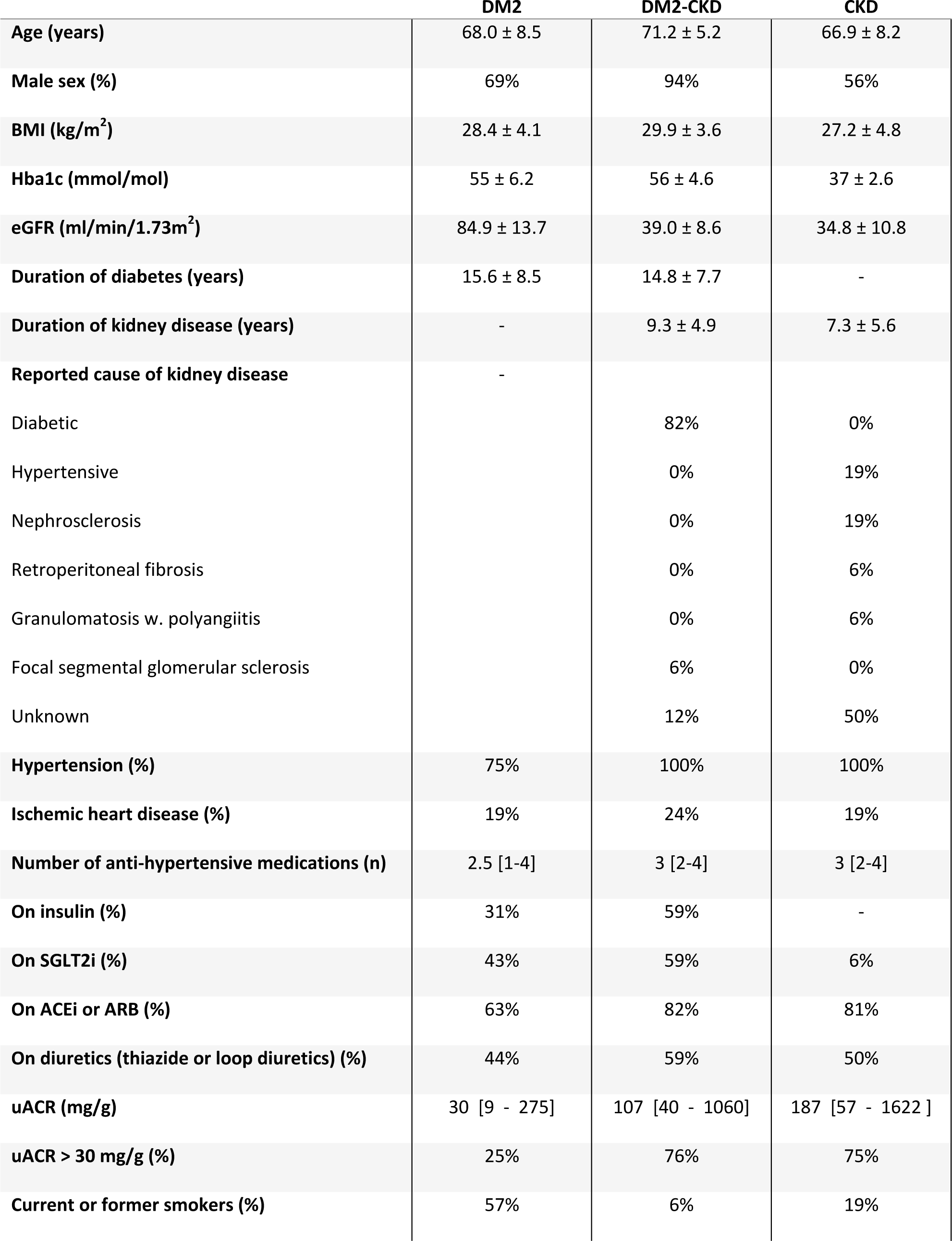
Baseline demographics. Data is presented as mean ± SD or median [interquartile range]. BMI: body mass index, HbA1c: glycated hemoglobin, eGFR: estimated glomerular filtration rate, uACR: urinary albumin/creatinine ratio, SGLT2i: sodium-glucose cotransporter 2 inhibitors, ACEi: angiotensin converting enzyme inhibitors, ARB: angiotensin receptor blockers.

|  | **DM2** | **DM2-CKD** | **CKD** |
| --- | --- | --- | --- |
| **Age (years)** | 68.0 ± 8.5 | 71.2 ± 5.2 | 66.9 ± 8.2 |
| **Male sex (%)** | 69% | 94% | 56% |
| **BMI (kg/m<sup>2</sup>)** | 28.4 ± 4.1 | 29.9 ± 3.6 | 27.2 ± 4.8 |
| **Hba1c (mmol/mol)** | 55 ± 6.2 | 56 ± 4.6 | 37 ± 2.6 |
| **eGFR (ml/min/1.73m<sup>2</sup>)** | 84.9 ± 13.7 | 39.0 ± 8.6 | 34.8 ± 10.8 |
| **Duration of diabetes (years)** | 15.6 ± 8.5 | 14.8 ± 7.7 | - |
| **Duration of kidney disease (years)** | - | 9.3 ± 4.9 | 7.3 ± 5.6 |
| **Reported cause of kidney disease** | - |  |  |
| Diabetic |  | 82% | 0% |
| Hypertensive |  | 0% | 19% |
| Nephrosclerosis |  | 0% | 19% |
| Retroperitoneal fibrosis |  | 0% | 6% |
| Granulomatosis w. polyangiitis |  | 0% | 6% |
| Focal segmental glomerular sclerosis |  | 6% | 0% |
| Unknown |  | 12% | 50% |
| **Hypertension (%)** | 75% | 100% | 100% |
| **Ischemic heart disease (%)** | 19% | 24% | 19% |
| **Number of anti-hypertensive medications (n)** | 2.5 [1-4] | 3 [2-4] | 3 [2-4] |
| **On insulin (%)** | 31% | 59% | - |
| **On SGLT2i (%)** | 43% | 59% | 6% |
| **On ACEi or ARB (%)** | 63% | 82% | 81% |
| **On diuretics (thiazide or loop diuretics) (%)** | 44% | 59% | 50% |
| **uACR (mg/g)** | 30 [9 - 275] | 107 [40 - 1060] | 187 [57 - 1622 ] |
| **uACR > 30 mg/g (%)** | 25% | 76% | 75% |
| **Current or former smokers (%)** | 57% | 6% | 19% |

### Renal Blood Flow

Due to poor image quality, two participants were excluded from the analysis, one in the DM2 group and one in the DM2-CKD group. In the DM2-CKD group empagliflozin reduced RBF by 6% (geometric mean ratio: 0.94, 95% CI: 0.91; 0.97, p < 0.001). There were non-significant reductions of 4% in the DM2-group and 1% in the CKD-group (p = 0.29 and 0.72). Changes in RBF were not significantly different between groups (p = 0.16). There was no specific period (p = 0.53) or carry-over effect (p = 0.82). The RBF results are presented in Table 2 and Figure 3.

**Figure 3:**
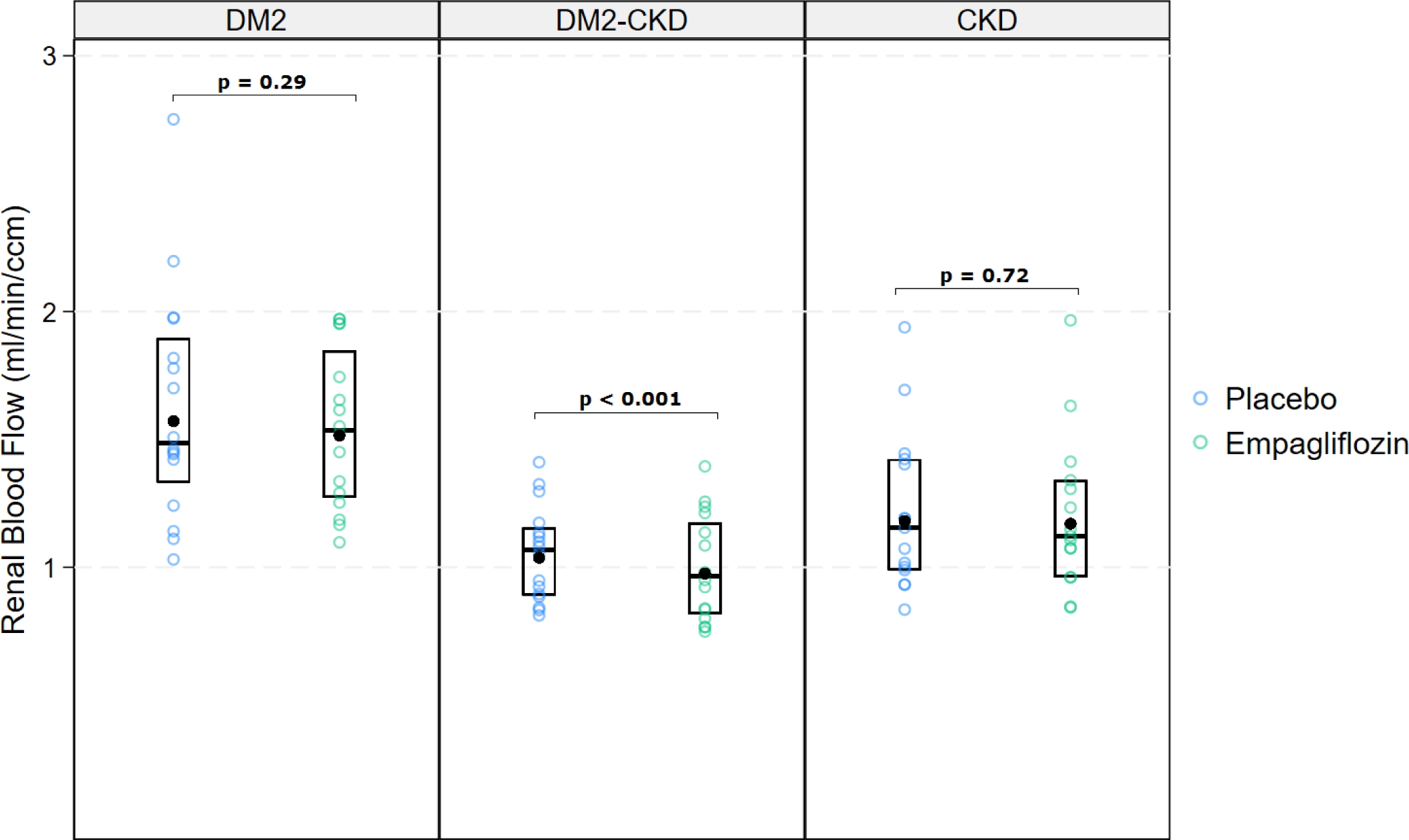
Renal blood flow. Median and interquartile range [IQR], black dot = geometric mean.

**Table 2:**
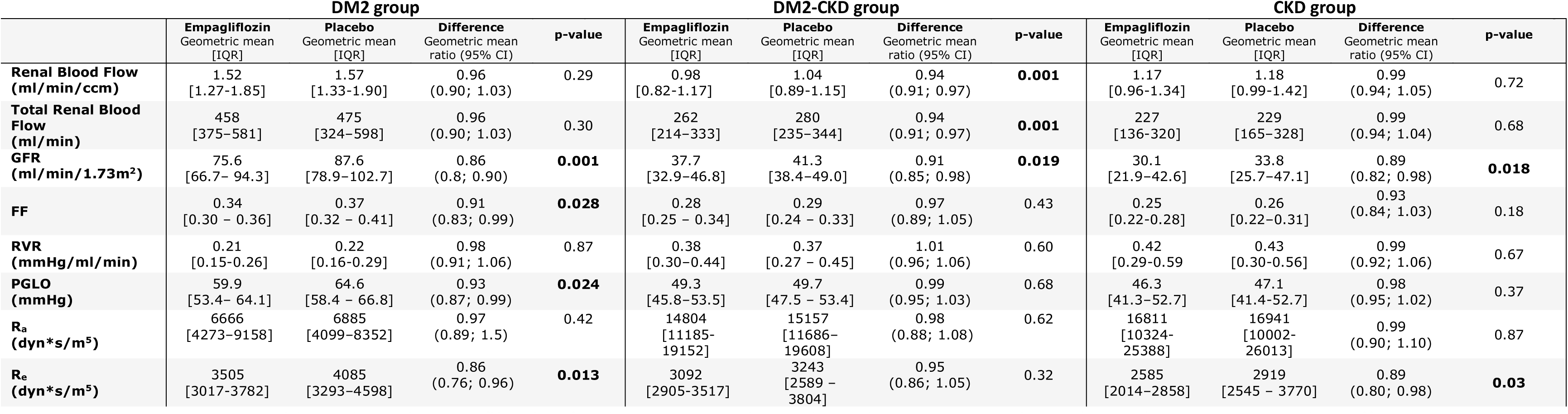
Renal hemodynamics. FF: filtration fraction, RVR: renal vascular resistance, PGLO: intra-glomerular pressure R_a_ and R_e_:afferent and efferent glomerular arteriolar resistance. P-values < 0.05 in bold.

### GFR

Empagliflozin significantly reduced GFR in all groups as compared to placebo. GFR was reduced by 14% in the DM2 group (p < 0.001), by 9 % in the DM2-CKD group (p = 0.019) and by 11% in the CKD group (p = 0.018). Changes between groups did not significantly differ (p = 0.36). There was no period or carry over effect (p = 0.98). GFR results are presented in Table 2 and Figure 4.

**Figure 4:**
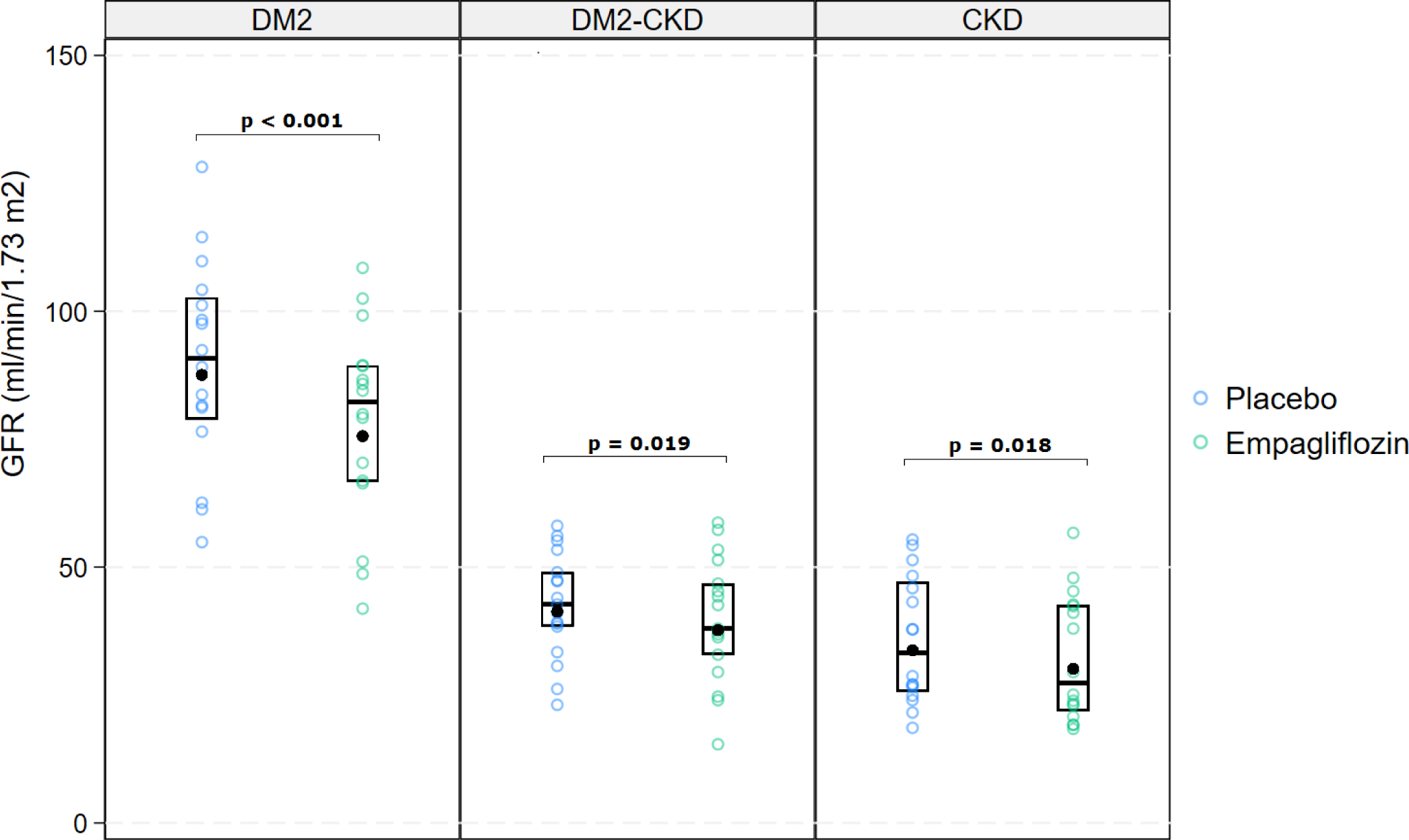
Glomerular filtration rate (GFR). Median and interquartile range [IQR], black dot = geometric mean.

### Systemic hemodynamics

Empagliflozin treatment reduced 24-hour systolic blood pressure by 5 mmHg in the DM2 group (95%CI: −9; −1 mmHg, p = 0.015), as compared to placebo. The mean reduction was 7 mmHg (95%CI: −12; −2 mmHg, p = 0.0097) in the DM2-CKD group and 4 mmHg in the CKD-group (95%CI: −9; 0 mmHg, p = 0.066). Empagliflozin lowered TVR in all groups. There was no period or carry over effects (p = 0.70 and 0.79). Heart rate did not change. One patient from the DM2 groups was excluded from this analysis due to missing data. See Table 3 and Figure 5.

**Figure 5:**
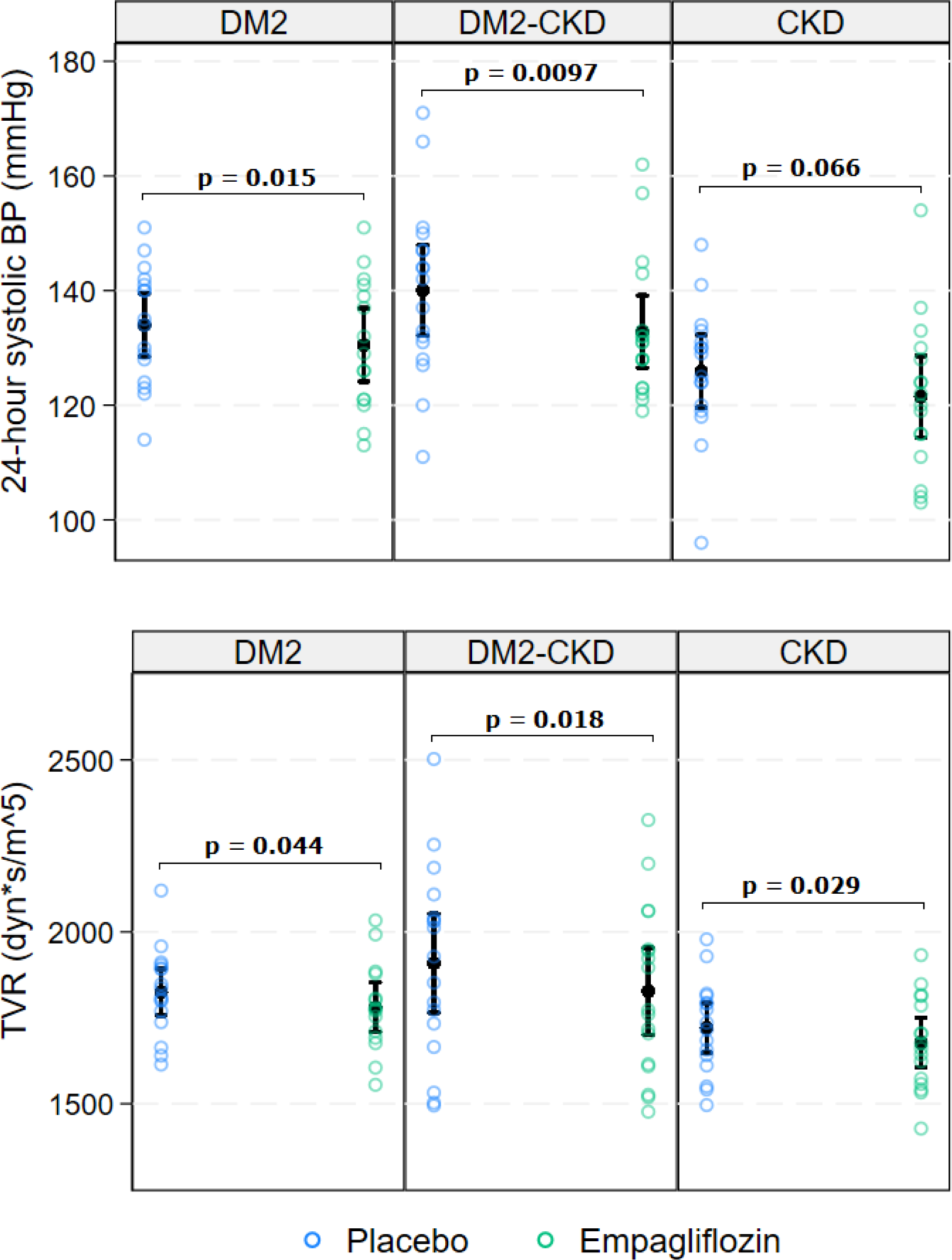
24-hour systolic blood pressure (BP), and total vascular resistance (TVR) with mean and 95% confidence interval (95%CI).

**Table 3:**
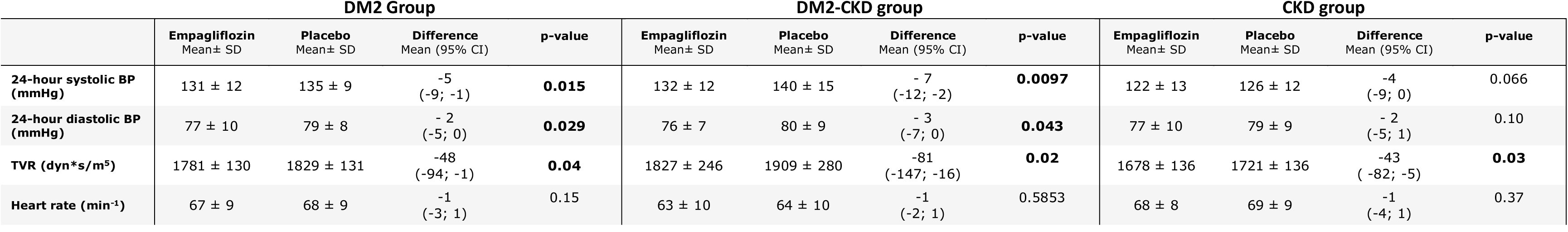
Systemic hemodynamics. TVR: total vascular resistance. P-values < 0.05 in bold.

### Renal hemodynamics

RVR did not change during empagliflozin treatment in any of the groups. FF was reduced by 9% in the DM2 group (p = 0.028), with no detectable change in the DM2-CKD or CKD groups (p = 0.42 and 0.72, respectively). P_GLO_ significantly decreased in the DM2 group with no change in the two other groups. R_e_ decreased in the DM2 and CKD groups. R_a_ did not change in any of the groups. Results are presented in Table 2.

### Changes in glucose and albuminuria

Analysis of uACR only included participants with an uACR of at least 30mg/g at the placebo visit. Empagliflozin significantly reduced uACR by 49% in the DM2-CKD group (p = 0.014). Although uACR was reduced by 50% in the DM2 group and 20% in the CKD group, these changes did not reach statistical significance (p =0.07 and p = 0.24, respectively). HbA1c did not change between treatments. Urinary glucose excretion was increased in all groups during empagliflozin treatment compared to placebo. See Table S2.

### Adverse events

The 58 participants who underwent randomization were evaluated for adverse events (AEs). AEs were classified as either related to empagliflozin or placebo treatment. If an AE happened during wash-out, it was ascribed to the previous treatment. 14 participants experienced an AE while treated with empagliflozin, while 19 participants experienced an AE on placebo. Details are presented in Table S1. Two participants dropped out due to AEs, one due to vaginal candidiasis during empagliflozin treatment and one due to an episode of dehydration and worsening of renal function during placebo treatment. One participant experienced a serious adverse event (SAE) of severe erysipelas requiring hospitalization and intra-venous antibiotics. This happened during wash-out after placebo treatment. One participant underwent an elective amputation of a toe due to a pre-existing chronic ulcer present before study enrolment. The amputation occurred during placebo treatment.

## Discussion

Our main finding is that empagliflozin reduced RBF in patients with DM2 and concomitant CKD. There was a smaller, non-significant decrease in the DM2 group and no change in the CKD group. This is the first time SGLT2i have been shown to affect RBF in patients with DM2. Furthermore, empagliflozin decreased GFR in all groups, reflecting the known acute reduction. [11, 22]. Our findings support the hypothesis that SGLT2i alleviate hyperfiltration in diabetic nephropathy. While, per definition, no participants in the DM2-CKD group had hyperfiltration defined as elevated GFR, it could still be present at the single nephron level [23] and the decrease in RBF and GFR we observe could indicate that empagliflozin reverses the TGF mediated feedback response, normalizes afferent arteriolar tone and diminishes hyperfiltration.

Another important finding is that empagliflozin decreased TVR in all groups, regardless of DM2 or CKD status. That SGLT2i decreases TVR has been reported in DM1, but to our knowledge never in DM2 or CKD [24]. These findings suggest a novel anti-hypertensive mechanism, perhaps related to a direct vasodilatory effect [25, 26]. Furthermore, we observed a clinically meaningful decrease in ambulatory BP in all groups during empagliflozin treatment in line with the existing evidence [27]. The lack of statistical significance in the CKD group probably reflects a small sample size rather than an absence of effect in this population.

RBF decreased significantly in the DM2-CKD group only. Between-group differences should be interpreted cautiously as the project was not designed for such comparisons, and while there was no statistical difference in RBF changes between groups, the change seems most pronounced in the DM2-CKD group. Diabetic kidney disease is characterized by overexpression and over activation of SGLT2 [28, 29] and sodium reabsorption - the main driver of TGF - increases during episodes of hyperglycaemia [30, 31]. Our findings suggest that individuals with diabetes and CKD experience a more prominent TGF response to SGLT2i. It is noteworthy that while the relative effects of SLGT2i on renal outcomes are similar across different patient categories [32], there is greater absolute treatment benefit in DM2 patients with both albuminuria and reduced eGFR [33]. Interestingly, it is also in patients with both DM2 and CKD we observe a marked decrease in RBF. However, as we examined only short-term effects, we cannot say if the changes we describe persist during chronic treatment or how they relate to the long-term benefits.

In the CKD group RBF did not change. The hemodynamic effects of SGLT2i have not previously been examined in patients with non-diabetic CKD and though our limited sample size prevents us from reaching a clear conclusion, our results could indicate that SGLT2 mediated changes in RBF could be less pronounced non-diabetic CKD than in diabetic nephropathy. Similarly, SGLT2i induce TGF responses less consistently in animal models with non-diabetic CKD than in those with diabetic nephropathy [34].

GFR decreased in all groups, while RBF decreased significantly in the DM2-CKD group only, suggesting GFR changes are not solely dependent on RBF. While identifying the exact mechanisms by which SGLT2i impact GFR is beyond the scope of this paper, several potential explanations could be considered. One possibility relates to the interplay between afferent- and efferent arteriolar resistance. While GFR and RBF respond in parallel to changes in afferent arteriolar resistance, changes in efferent resistance can dissociate autoregulation of GFR from RBF [35]. It has previously been suggested that SGLT2i can alleviate hyperfiltration through dilation of the efferent arteriole, eliciting a decrease in GFR without affecting RBF [36]. However, GFR is not determined solely by glomerular pressure; intra-tubular pressure and properties of the glomerular filtration barrier could also be of importance. By inhibiting solute reabsorption from the distal tubule, SGLT2i could theoretically increase tubular pressure leading to reduced pressure gradient across the glomeruli and a decrease in GFR [37]. Additionally, animal studies have shown that dapagliflozin can directly affect podocyte function [38]. SGLT2i could thus potentially exert non-hemodynamic effects on GFR.

We calculated estimates of intra-renal hemodynamics to further explore the hemodynamic effects of SGLT2i. These calculations are indirect and can at best be indicative of the underlying physiology; the Gomez equations rely on assumptions of normal renal physiology which do not align with the characteristics of our cohort, particularly those with CKD. Furthermore, potential non-hemodynamic effects on GFR are not taken into account [21]. Nevertheless, as more direct methods in humans are lacking, they provide our best estimates. Using these estimates, we found that empagliflozin decreased FF, P_GLO_ and R_e_ in the DM2 group, which is in agreement with previous findings from van Bommel et al. who studied a comparable cohort of patients with DM2 and preserved kidney function [15]. This supports the current view that SGLT2i could exert effect in DM2 partly through dilation of the efferent glomerular arteriole. We saw a decrease in R_e_ in the CKD group as well, perhaps indicative of a similar mechanism.

Despite significant changes in RBF in the DM2-CKD group, we observed no change in RVR in this or any other group, nor did R_a_ change. This seemingly contrasts the results from Cherney et al., who found that empagliflozin decreased ERPF and increased RVR in patients with DM1 and hyperfiltration, indicating an increase in pre-glomerular resistance [7]. We do not, however, believe our findings preclude an SGLT2i induced effect the afferent arteriole. While R_a_ did not change, TVR decreased significantly. This is noteworthy, since we would have expected a parallel auto regulatory decrease in R_a_ to conserve RBF as has previously been described with for example angiotensin receptor blockers [39]. That R_a_ did not decrease while RBF did suggests a SLGT2i-induced relative afferent vasoconstriction. Our findings thus indicate a combined afferent and efferent arteriolar response. Similar mechanisms were described in a study in diabetic rats [40].

We measured RBF using a novel ^82^Rb-PET/CT method. The method has low intra-assay variability and excellent inter-observer reliability, allowing for precise and reliable measurements of relative changes in perfusion [20]. When studying SGLT2i, it has the further advantage compared to para-amino-hippuric acid (PAH) of not being affected by glycosuria [41]. There are disadvantages to the method as well; ^82^Rb-PET/CT likely underestimates perfusion at high perfusion rates [15]and the method cannot distinguish between cortical and medullar perfusion [19, 42]. Furthermore, ^82^Rb is a potassium analogue and thus not an inert tracer and we have not taken potential urinary excretion into account [42]. Altogether, ^82^Rb-PET/CT likely underestimates true RBF, which can explain why the measured values are lower than what have previously been described [15], in a cross over design, absolute values are of less importance as we examine changes within the same individual. Our project has other limitation; the small sample sizes make comparisons between groups difficult. Furthermore, changes in RBF were smaller and variability larger than anticipated in the power calculation, so we cannot rule out potential effects in the DM2 and CKD-groups had the sample sizes been larger. Inclusion in the DM2-CKD group did not require a biopsy verified diagnosis of diabetic nephropathy, so other aetiologies of CKD could be present. Likewise, in the CKD-group the cause of CKD was not reported in half the participants, limiting generalizability. We did not employ glycaemic clamping method nor were participants fasting. While this may add uncertainty to the results, an advantage is that our approach more accurately reflects both the patient populations and conditions in which SGLT2i would be used in a real-world setting.

In conclusion, short-term empagliflozin treatment significantly reduces RBF in patients with DM2 and concomitant CKD as compared to placebo, while GFR, BP and TVR was reduced in all groups. Our findings suggest that the renal hemodynamic effects of SGLT2i could be caused by relative pre-glomerular vasoconstriction, as well as post-glomerular vasodilation.

## Data Availability

All study data will be made available upon request.

## Authors contribution

All authors contributed to the manuscript. SFN, FHM, NHB and JNB designed the trials, which were conducted by SFN with support from CLD and FHM. SFN analyzed the data and interpretation was done in cooperation with FHM, JNB and NHB. SFN drafted the manuscript which was revised and accepted by all authors.

## Acknowledgements

We would like to thank the biomedical laboratory scientists at The University Clinic in Nephrology and Hypertension and The Department of Nuclear Medicine, Gødstrup for making this project possible. Special thanks to Lene Ring Madsen and STENO Diabetes Center, Aarhus for helping with recruitment in the DM2-CKD group and to Stine Sundgaard Langaa for kindly helping with the analysis of the Rb^82^-PET/CT scans.

## Data sharing

All study data will be made available upon request.

## Non-standard abbreviations and acronyms

82Rb-PET/CT: ^82^Rubidium positron emission tomography/computed tomography
99mTc-DTPA: ^99m^Technetium-diethylene-triamne-pentaacetate
AA: abdominal aorta
ADPKD: autosomal dominant polycystic kidney disease
AE: adverse event
BMI: body mass index
BP: blood pressure
CKD: chronic kidney disease
DM1: type 1 diabetes mellitus
DM2: type 2 diabetes mellitus
eGFR: estimated glomerular filtration rate
ERFP: effective renal plasma flow
EVF: erythrocyte volume fraction
FF: filtration fraction
GCP: good clinical practice
GFR: glomerular filtration rate
HbA1c: glycated hemoglobin
IQR: inter quartile range
MAP: mean arteriel pressure
P_GLO_: intra glomerular pressure
R_a_: afferent glomerular arteriolar resistance
RBF: renal blood flow
R_e_: efferent glomerular arteriolar resistance
RVR: renal vascular resistance
SAE: serious adverse event
SD: standard deviation
SGLT2i: sodium-glucose cotransporter 2 inhibitors
TGF: tubulo-glomerular feedback
TVR: total peripheral vascular resistance
uACR: urinary albumin/creatinine ratio
VOI: volume of interest

---

**Figure S1.**
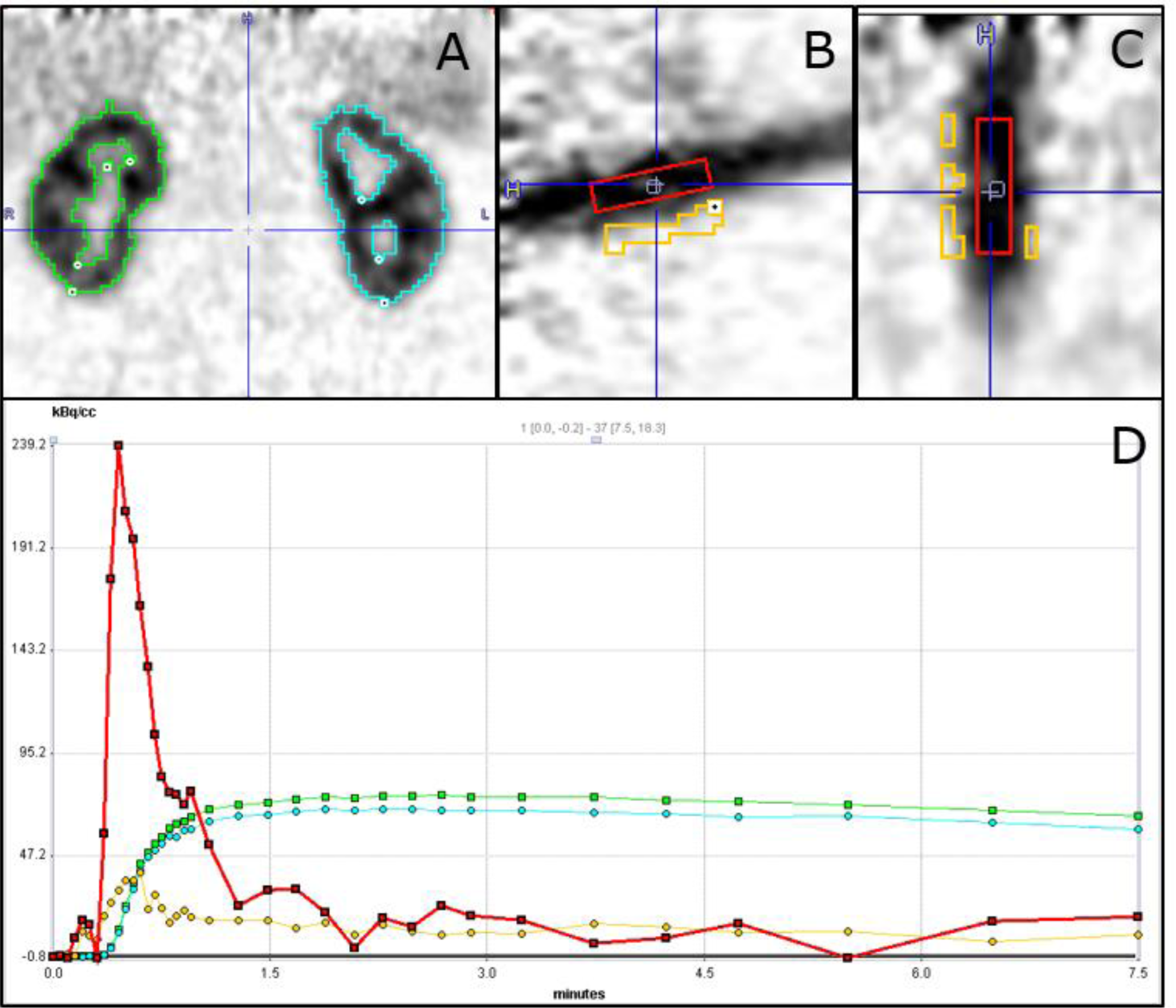
^82^Rb-PET/CT renal scan. **A:** Renal activity of ^82^Rb with iso-contours of the right and left kidney (green and turquoise), coronal view. **B and C:** The abdominal aorta (AA) with volume of interest (VOI) (red) and a background box for partial volume correction (orange), sagittal and coronal views. **D:** Time activity curves of ^82^Rb activity in the AA (red), right and left kidney (green and turquoise) and AA background (orange)

**Table S1:** Adverse events

| Event | Empagliflozin | Placebo |
| --- | --- | --- |
| Number of participants with an adverse event: | 14 | 19 |
| Hypoglycemia | 1 | 2 |
| Musculoskeletal | 2 | 6 |
| Urogenital infections | 2 | 1 |
| Dizziness | 2 | 3 |
| Headache | 0 | 1 |
| Infection | 2 | 2 |
| Fatigue | 1 | 0 |
| Amputation (toe) | 0 | 1 |
| Hyperglycemia | 0 | 1 |
| Polyuria | 1 | 0 |
| Diarrhea | 1 | 1 |
| Dehydration | 0 | 1 |
| Worsening of renal function | 0 | 1 |
| Hyponatremia | 0 | 1 |
| Other | 5 | 4 |

**Table S2:** Glycated hemoglobin (HbA1c), plasma glucose, albumin/creatinine ratio and urinary glucose. P-values < 0.05 in bold. * Only participants with an uACR > 30 mg/g at the placebo visit. N = 4 in the DM2 group. N = 14 in the DM2-CKD group. N = 11 in the CKD group

|  |  |  |  |  |  |  |  |  |  |  |  |  |
| --- | --- | --- | --- | --- | --- | --- | --- | --- | --- | --- | --- | --- |
| **Plasma glucose (mmol/L)** | 9.5 ± 2.1 | 11.1 ± 4.2 | -1.6<br>(-3.4; 0.2) | p = 0.078 | 9.4 ± 2.4 | 9.6 ± 2.8 | -0.1<br>(-1.2; 1.0) | P = 0.81 | 5.6 ± 1.1 | 5.9 ± 0.9 | -0.2<br>(-0.7; 0.3) | P = 0.32 |
|  | **Empagliflozin**<br>Geometric mean<br>[IQR] | **Placebo**<br>Geometric mean<br>[IQR] | **Difference**<br>Geometric mean ratio<br>(95% CI) | **p-value** | **Empagliflozin**<br>Geometric mean<br>[IQR] | **Placebo**<br>Geometric mean<br>[IQR] | **Difference**<br>Geometric mean ratio (95% CI) | **p-value** | **Empagliflozin**<br>Geometric mean<br>[IQR] | **Placebo**<br>Geometric mean<br>[IQR] | **Difference**<br>Geometric mean ratio (95% CI) | **p-value** |
| **Albumin/creatinine ratio\* (mg/g)** | 347<br>[133–1087] | 694<br>[286–2063] | 0.50<br>(0.22; 1.12) | p = 0.07 | 205<br>[68–644] | 399<br>[136 – 1326] | 0.51<br>(0.31; 0.85) | **P = 0.014** | 280<br>[57–1332] | 350<br>[80–1921] | 0.80<br>(0.54; 1.29) | p = 0.24 |
| **Urinary glucose (mmol/L)** | 87.9<br>[53.3 – 138.4] | 0.2<br>[0.0–1.6] | - | **p < 0.001** | 41.1<br>[38.0–66.2] | 0.1<br>[0.0–0.1] | - | **p < 0.001** | 11.6<br>[7.3–26.1] | 0.0<br>[0.0–0.0] | - | **p < 0.001** |

